# Stakeholder’s perspective on Brain-Computer Interfaces for children and adolescents with quadriplegic cerebral palsy

**DOI:** 10.1101/2024.05.23.24307809

**Authors:** M.P. Branco, M.S.W. Verberne, B.J. van Balen, A. Bekius, S. Leinders, M. Ketelaar, J. Geytenbeek, M. van Driel-Boerrigter, M. Willems-op het Veld, K. Rabbie-Baauw, M.J. Vansteensel

**Affiliations:** UMC Utrecht Brain Center, Department of Neurology and Neurosurgery, Utrecht, The Netherlands; Human Technology Interaction, Eindhoven University of Technology, Eindhoven, The Netherlands; Ethics and Philosophy of Technology, Delft University of Technology, The Netherlands; Center of Excellence for Rehabilitation Medicine, UMC Utrecht Brain Center, University Medical Center Utrecht, and De Hoogstraat Rehabilitation, Utrecht, The Netherlands; UMC Amsterdam, Department of Rehabilitation medicine, CP expertise Center, Amsterdam, The Netherlands; Patient Association CP Nederland, Houten, The Netherlands; Patient representative, Baauwopmij, The Netherlands

**Keywords:** survey, user-centered design, brain-computer interface, communication, cerebral palsy, children

## Abstract

Communication Brain-Computer Interfaces (cBCIs) use neural signals to control a computer and are of interest as a communication tool for people with motor and speech impairment. Whereas the majority of cBCI research focuses on adults, the technology may also benefit children and adolescents with communication impairments, for example as a result of cerebral palsy (CP). Here we aimed to create a solid basis for the user centered design of cBCIs for children and adolescents with CP and complex communication needs by investigating the perspectives of their parents/caregivers and health care professionals on communication and cBCIs. We conducted an online survey on 1) current communication problems and usability of used aids, 2) interest in cBCIs, and 3) preference for specific types of cBCIs. A total of 19 parents/caregivers and 36 health care professionals who live(d) or work(ed) directly with children and adolescents (8-25 years old) with quadriplegic CP participated. Both groups of respondents indicated that, of 12 potential communication-limiting factors, motor impairment occurred the most frequently and also had the greatest impact on communication. The currently used communication aids included mainly no/low-tech aids (e.g., letter card) and high-tech aids (e.g., tablet or computer). Mid-tech aids (e.g., systems with static displays) were less frequently used. The majority of health care professionals and parents/caregivers reported an interest in cBCIs for children and adolescents with severe CP, with a slight preference for implanted electrodes over non-implanted ones, and no preference for either of the two proposed mental BCI control strategies (visual stimuli and imagined/attempted movement). These results indicate that cBCIs should be considered for a subpopulation of children and adolescents with severe CP, and that in the development of cBCIs for this group both P300 and sensorimotor rhythms, as well as the use of implanted electrodes, should be considered.

## 1. Introduction

Communication during childhood is essential to acquire receptive and expressive language skills **(Colonnesi et al. 2010; D’souza, D’souza, and Karmiloff-Smith 2017)** and to be able to effectively interact with others **(Kastner, May, and Hildman 2001; Hohm et al. 2007)**. However, for children affected by a form of physical impairment such as cerebral palsy (CP) **(Bax et al. 2005; Odding, Roebroeck, and Stam 2006; Gulati and Sondhi 2018)** effective communication is not always self-evident. In Europe alone, approximately 10.000 children are born with CP each year **(Andersen et al. 2008; Oskoui et al. 2013; Himmelmann and Uvebrant 2018; McIntyre et al. 2022)**, and worldwide over 17 million individuals live with this physical disability according to the Cerebral Palsy foundation (https://www.yourcpf.org/statistics/) and the Cerebral palsy Alliance Research Foundation (https://cparf.org/what-is-cerebral-palsy/facts-about-cerebral-palsy/). CP is a group of disorders, caused by a range of non-progressive disturbances in the immature brain occurring before, during or shortly after birth, that permanently affect the ability to, among others, move and communicate **(Rosenbaum et al. 2007)**. In particular, children with bilateral spastic CP and dyskinetic CP **(Himmelmann et al. 2009; Shevell, Dagenais, and Hall 2009)** are among the most severely affected. Motor functioning of these children is typically classified by Gross Motor Function Classification System (GMFCS; **Palisano et al. 1997**) and Manual Ability Classification System (MACS; **Eliasson et al. 2006**) level IV or V (hereafter referred to as quadriplegic CP or quadCP). A significant percentage of these children has a normal or close-to-normal IQ and age-appropriate language comprehension skills **(Andersen et al. 2008; Geytenbeek et al. 2015; Reid et al. 2018)**. Yet, because of their motor impairment, they are unable to communicate effectively **(Parkes et al. 2010; Mei et al. 2020)**, which negatively affects their educational development and social interactions. Providing a means to establish effective communication would have a direct impact on the child’s development and ultimate societal participation **(Hetzroni 2004; Voorman et al. 2010; Light and McNaughton 2011; Coleman et al. 2013)**.

For children and adolescents with quadCP, the use of augmentative and alternative communication (AAC) aids is essential for communication **(Marshall and Goldbart 2008)**. Several AAC technologies are available to facilitate communication **(Griffiths and Addison 2017; Cooley Hidecker 2020)**, which all rely on some level of reliable muscle control (e.g., hand or eye movement) for its use. A communication Brain-Computer Interface or cBCI **(Vansteensel and Jarosiewicz 2020)** could be a completely novel, and perhaps ideal AAC solution for children with quadCP who have difficulty using conventional AAC. A cBCI uses brain signals to directly control communication software, without the need for any muscle control **(Wolpaw and Wolpaw 2012)**. Despite the great progress in the development of cBCIs, and promising results in demonstrating cBCI-based communication in adults with severe motor impairment **(Vansteensel et al. 2016; Oxley et al. 2021; Moses et al. 2021; Metzger et al. 2023; Willett et al. 2021)**, the pediatric application of this technology has been left mostly untouched **(Kinney-Lang, Auyeung, and Escudero 2016; Orlandi et al. 2021; Bergeron et al. 2023)**. This situation provides an excellent opportunity to adopt a user centered design approach for the exploration and development of cBCIs for children and adolescents with quadCP. A user centered design places the user and their needs at the center throughout all stages of the development process **(Kübler et al. 2014)**. An important aspect of this approach is the generation of an in-depth understanding of the user and the context of use. Initial efforts to involve the young population with CP have focused on assessing the awareness about BCIs within the CP community **(Letourneau et al. 2020)**, and determining the subpopulation of children with CP that could benefit from BCIs . Yet, it is still unclear how cBCIs can benefit individuals with quadCP beyond current AAC solutions and what the need and wishes of this population are regarding cBCIs. Moreover, it is unclear if there is a preference of this target population for a specific neural signal recording method **(Leuthardt, Moran, and Mullen 2021)** or for a specific mental strategy (e.g., motor imagery/attempt or visual stimuli) to accomplish cBCI control **(Branco et al. 2021)**. It is, therefore, crucial to further map the needs and interests of the target population in order to design the best clinically viable cBCI system for children and adolescents with quadCP.

In this study we mapped the needs and wishes regarding cBCIs as an AAC technology for children and adolescents (8-25 years old) with quadCP in the Netherlands. For that purpose, we assessed the perspective of two relevant stakeholder populations related to this group: parents/caregivers and health care professionals. We performed a detailed online survey that focused on three main questions being 1) what communication aid children and adolescents currently use and how well these tools serve their needs; 2) the interest in cBCIs as an AAC solution; and 3) the preferences for the type of cBCI (implanted versus non-implanted) and mental cBCI control strategy.

## 2. Material and Methods

An online survey was administered to parents/caregivers and health care professionals who live(d) or work(ed) directly with children and adolescents with quadCP (GMFCS level IV and V). The surveys were written in Dutch and were implemented, distributed and collected in Qualtrics (www.qualtrics.com). The study was assessed by the medical research ethics committee of the Utrecht Medical University Center, who deemed this study exempt from the Dutch Medical Scientific Research Act (non-WMO). Survey description and links were posted on the website of the patient organization CP Nederland (https://cpnederland.nl) and shared via the social media channels of CP Nederland, researchers and user advocates involved in this study. Participants gave explicit informed consent to participate in the research and for the use of their data on the first page of the survey, and by completing and submitting the survey.

### 2.1 Participants

#### 2.1.1 Health care professionals

With this survey we aimed to reach Dutch health care professionals who previously or currently worked with children and adolescents with quadCP, such as speech therapists, physiotherapists or occupational therapists. In total, 69 health care professionals showed interest in taking part in the research, of which 36 were included in this study (52% inclusion rate). Reasons for exclusion included incomplete survey (if the expiration date was exceeded or they did not work with the quadCP target-group with GMFCS IV-V) and incorrect population (if they did complete the survey but indicated in the survey not being a care professional). Two versions of the survey were deployed, which only differed in the need for pre-registration. The first version required pre-registration using a form. The link to the survey was emailed to the individuals who registered, to avoid spam and bot replies. The second version used a public link to the survey, to facilitate participation. Both versions of the survey were completed by 18 participants. Survey responses were stored anonymized and coded for all participants.

#### 2.1.2 Parents/caregivers

In addition, we aimed to reach parents and caregivers (for example legal guardians or other non-professional caregivers) of children and adolescents with quadCP. Parents/caregivers expressed interest through a pre-registration form, after which a survey link was sent via email. In total, 25 parents/caregivers completed the pre-registration form, of which 19 were included (76% inclusion rate). Reasons for exclusion were incomplete survey (due to exceeded expiration date), incorrect age-group of their child with CP (younger than 8 years old or older than 25 years old) and incorrect level of impairment of their child with CP (GMFCS level I-III, or if their child was verbally intelligible for strangers). Given that the GMFCS term may not be familiar for some parents/caregivers (2 out of 19 parents/caregivers reported GMFCS of ‘unknown’), an additional question about verbal intelligibility for strangers (‘Can your child speak clearly to strangers?’) was used to further assess eligibility.

### 2.2 Structure of the questionnaire

The online survey consisted of four parts: 1) information letter and informed consent, 2) screening and demographics, 3) communication barriers and currently used communication AAC, 4) interest in cBCIs and types of cBCIs. Animation videos narrated in Dutch were used to explain the concept of cBCI and the types of cBCIs in layman terms, similar to previous studies **(Branco et al. 2021; 2023)**.

#### 2.2.1 Information letter and informed consent

The goal and duration (approximately 20 minutes) of the online questionnaire was described in a participant information letter. Health care professionals participating in the first survey version and all parents/caregivers received the participant information letter by email. The introductory text of the survey referred to the letter. For health care professionals participating in the second survey version, a link to the letter was provided at the start of the survey. Participants were asked to give explicit (yes/no) informed consent to participate in this research and for the use of their data for this study.

#### 2.2.2 Screening and demographics

Screening questions to confirm inclusion criteria of the study, including age and GMFCS level/verbal intelligibility of the child(ren) with CP they cared for, and the nature of the relation with this group (either work or family) were asked using multiple-choice questions in the first part of the survey. In addition, general demographics of the participants (e.g., age, gender, education) and their child/client target-group (e.g., type(s) of CP and, for parents/caregivers, gender of their child) were asked using multiple-choice or short-answer questions.

#### 2.2.3 Communication barriers and current AAC

In the second part of the survey, we asked about the AAC aids the children/clients of the participants mostly used, and about the most common barriers in communication, such as problems with moving, hearing or seeing, or problems with usage or functionality of the used communication aids. We divided AAC aids into three categories (www.isaac-nf.nl/communicatiehulpmiddelen/), namely ‘Cat 1’ – no- or low-tech communication, without AAC aids or with a simple AAC aid (e.g., lettercard); ‘Cat 2’ – mid-tech communication, through simple speech-computer devices (e.g., static systems); or ‘Cat 3’ – high-tech communication with AAC aids (dynamic systems, including tablet or computer). For each category we identified where, with whom, and for what goal these aids were used, and assessed the level of user satisfaction on a 5-point Likert scale and via open-ended questions on the advantages and disadvantages of each selected aid.

#### 2.2.4 Need for cBCIs and types of cBCIs

The last section of the survey focused on BCIs for communication. First, we asked the participants if they were acquainted with the concept of a BCI before this research and if so, what their impression/opinion about BCIs was until now (5-point Likert scale). Second, we introduced the concept of cBCI by means of an animation video (duration 81 seconds) followed by a question of the participant’s opinion about usefulness of cBCIs for the quadCP target-group. Third, we introduced two types of mental strategies to control a cBCI, namely using visual stimuli (i.e., P300 or steady-state visual evoked potentials) and motor attempt (i.e., attempted or imagined movement of the hand), as well as two techniques to measure brain signals, namely scalp electroencephalography (EEG) and implanted electrodes (i.e., intracranial electrodes), using two animation videos (duration 70 seconds and 82 seconds, respectively). In the videos we described EEG as electrodes placed on the outside of the head, which require precise placement often with the use of gel, and the use of which may not be possible or comfortable in certain situations, such as during sleep. We described implanted electrodes as electrodes placed in the head (for example on the surface of the brain) by means of surgery and opening of the skull, which involves surgical risks. Implanted electrodes were described to be available 24 hours per day and esthetically invisible to others. No referral was made to the level of effectiveness or efficiency of BCI control that could be accomplished with the two techniques. For each item, participants were asked to rate their applicability for children and adolescents with quadCP using a 5-point Likert scale. After every rating question the participants had the chance to add comments and suggestions through free text fields.

### 2.3 Statistical analysis

#### 2.3.1 Quantitative analysis

Multiple-choice and closed- and short-answer questions were described using descriptive statistics. Frequencies or percentages relative to the total number of respondents were used to compare results between the two groups (health care professionals and parents/caregivers). Responses to ranking questions were quantified using the center-of-mass (COM) score as described in **(Vansteensel et al. 2017; Branco et al. 2023)**. This metric allows to summarize the overall ranking place of a category across respondents. The COM score was computed per group (health care professionals or parents/caregivers) depending on the number of ranks available to each group for a specific question. The lowest rank (e.g., least frequent or least impairing) were attributed a weight of 1 and the highest rank (e.g., most frequent or most impairing) a score corresponding to the number of total number of ranks (either 3, 4, 6 or 12). A weight of zero was used for null answers (e.g., 0% or ‘never’), answers not chosen or not applicable. Independent sample t-tests were used where appropriate to compare level of interest or satisfaction between or across groups.

#### 2.3.2 Qualitative analysis

Responses to open questions were analyzed using ‘content analysis’ **(Hsieh and Shannon 2005; Elo and Kyngäs 2008)** using NVivo software (www.lumivero.com). We used an inductive approach and derived codes directly from the text. Two of the authors (BVB and SL) coded the statements of the respondents independently in an iterative fashion and then agreed on a list of codes that covered all the issues described by respondents. Separate code-lists were generated per group of participants (health care professionals and parents/caregivers) and per open question if there were enough informative answers to draw a meaningful conclusion. After agreeing on an initial list of codes, all statements were annotated with one or more codes from that list by BVB and MV. After two rounds of updating codes and a third round of discussion with BVB, MV, and MJV, consensus was reached about code assignments. Where relevant, the codes were grouped into themes. In total, five open questions were analyzed for both groups of participants, namely about 1) experienced communication barriers, 2) wishes and desires (both questions from section 2.2.3 on ‘Communication barriers and current AAC’), 3) meanings and aims of BCIs, 4) mental strategies for cBCI-control, and 5) techniques to measure brain activity (3, 4, and 5 were answered based on section 2.2.4: ‘Need for cBCIs and types of cBCIs’). Qualitative analysis was fully performed in the native language of the respondents (in Dutch), and several respondents’ quotes were translated to English for the purpose of this manuscript.

## 3. Results

### 3.1 Demographics

We included 36 health care professionals (median age 46 years old, range 26-65 years old; 94% female, 3% male and 3% unknown) with a diverse professional background, ranging from speech therapists (36%) to rehabilitation physicians (14%), and the majority (66%) having more than ten years of professional experience with children and adolescents with quadCP **(Figure 1A)**. In addition, we included 19 parents/caregivers (median age 49.5 years old, range 34 to 56 years old; 95% female, 5% male) of children with quadCP with an age range of 8-25 years old **(Figure 1B)**. While health care professionals typically worked with individuals with different types of CP and a wide range of GMFCS scores (I-V), parents/caregivers included in this study reported on their children with dyskinetic, spastic bilateral, or ataxic CP, and high GMFCS levels (IV and V) and who were not verbally intelligible to strangers **(Figure 1C)**.

**Figure 1.**
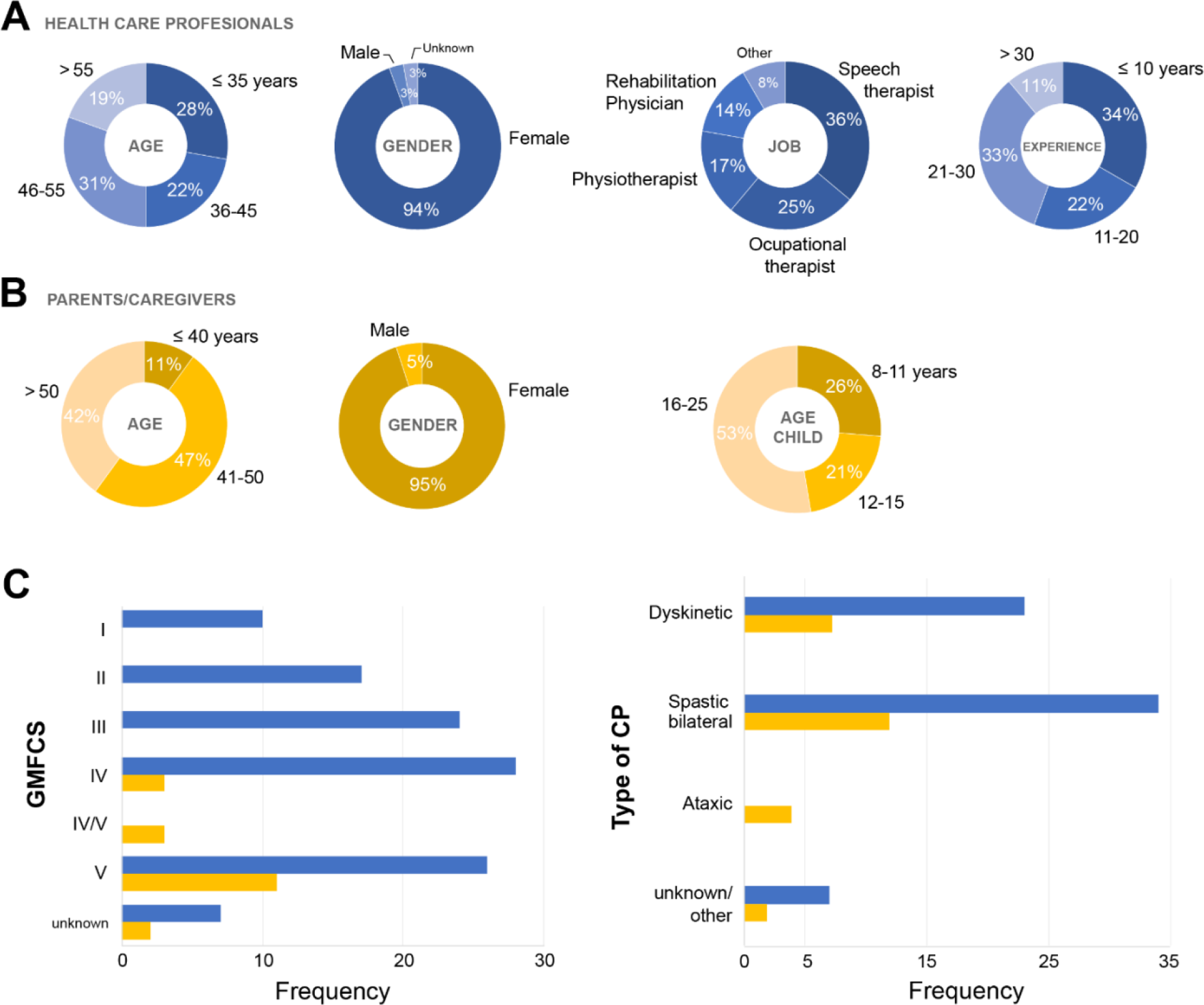
Demographic information of the health care professionals and parents/caregivers. (A-C) Health care professionals are indicated in blue and parents/caregivers in yellow. Demographics expressed in percentages (%) of (A) the health care professionals (N = 36) and (B) the parents/caregivers (N = 19), as extracted from part 1 of the survey. (A) Age (in years), gender, professional occupation (job; other=caregiver, special education teacher or childcare worker) and years of experience with the target quadCP group of the included health care professionals. (B) Participant age (in years) and gender as well as age of child/adolescent with quadCP (in years) of the included parents/caregivers. (C) Gross Motor Function Classification System (GMFCS) level (I-V) and type of CP of the children/clients of the respondents, expressed in frequencies of respondents (multiple answers possible).

### 3.2 Communication barriers and current AAC

When rating factors that generally affect communication, both health care professionals and parents/caregivers scored problems with movement (‘Motor’) as the most frequently occurring communication-impairing factor **(Figure 2A)** and also as the factor that impairs communication to the largest extent **(Figure 2B)**. Interestingly, health care professionals and parents/caregivers disagreed on importance of other factors, with health care professionals indicating problems with cognition and thinking ability (‘Cognition’) and problems with learning to read and spell (‘Read and spell’) more frequently occurring than parents/caregivers **(Figure 2A)**. On the other hand, parents/caregivers also rated problems related to the communication partner’s difficulty to follow the communication signals from the child as an often-occurring barrier **(Figure 2A)** that also greatly affects communication **(Figure 2B)**. Even though health care professionals reported problems with reading and spelling (‘Read and spell’) to occur often, they reported that this affects communication less than other factors such as understanding verbal language (‘Understanding verbal’) **(Figure 2B)**. The least prevalent and the least impairing factor reported by both groups was auditory problems (‘Auditory’).

**Figure 2.**
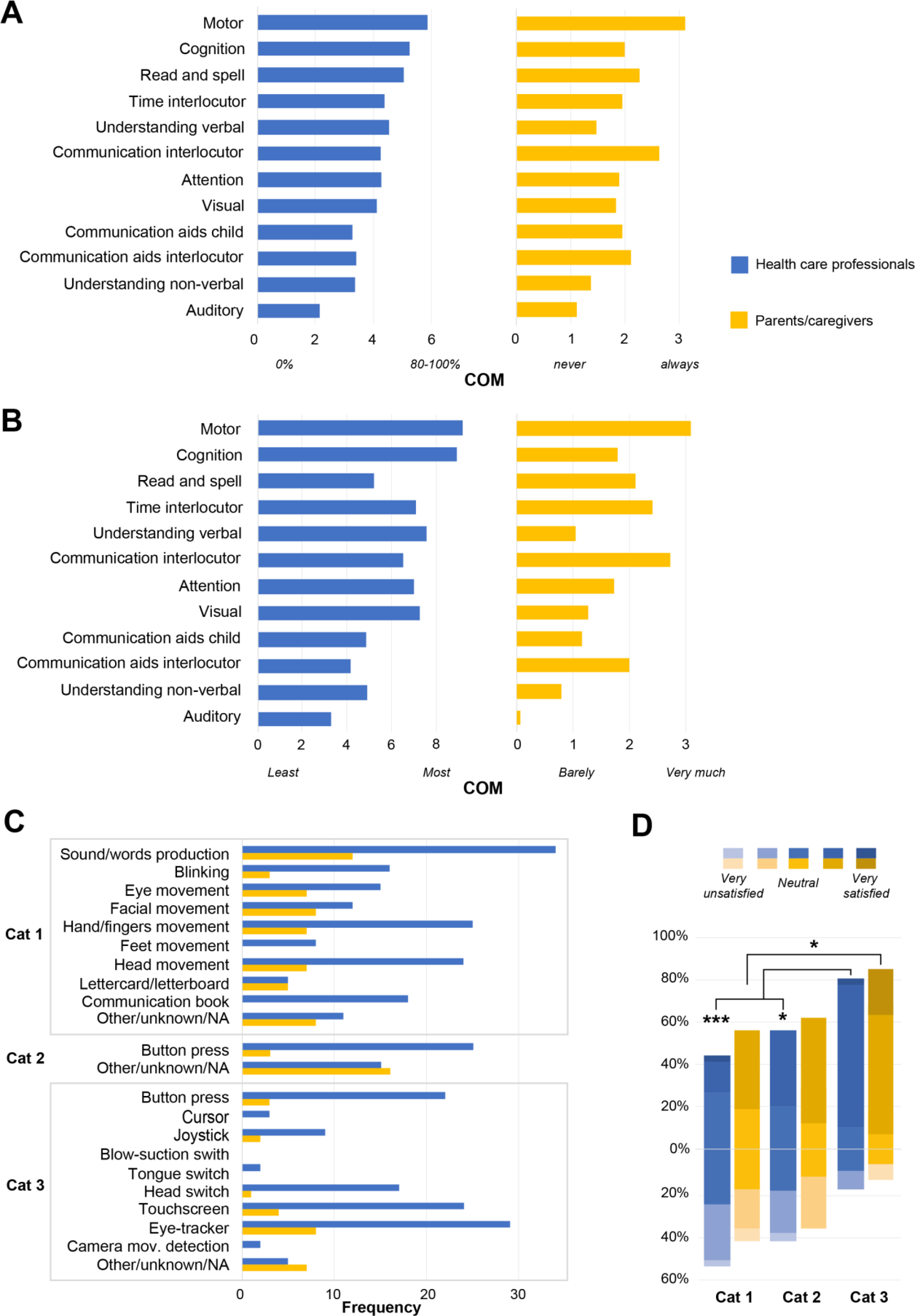
Communication barriers and current AAC. (A-D) Health care professionals are indicated in blue and parents/caregivers in yellow (A) Prevalence of communication barriers. Health care professionals indicated percentage of occurrence of each factor in their client group using 6 ranks (0%, 1-20%, 20-40%, 40-60%, 60-80% and 80%-100%), whereas parents/caregivers used 4 ranks (‘never’, ‘sometimes’, ‘often’ or ‘always’) to describe the occurrence of the respective factor in their child. (B) Indication of how much each barrier impairs communication of children and adolescents with quadCP. Health care professionals ranked each barrier from least (1) to most (12) impairing, whereas parents/caregivers used 4-rank scale (‘barely’, ‘a little’, ‘much’ and ‘very much’). (A-B) Ranks are indicated using a center- of-mass (COM) score, which ranges from 1 to a maximum number of ranks per group and question. (C) Frequency of respondents who report their child/client(s) using a type of AAC, divided into 3 categories: ‘Cat 1’ no- or low-tech, ‘Cat 2’ mid-tech, and ‘Cat 3’ high-tech aid. (D) Percentage of respondents who rated the level of satisfaction of each AAC category using a 5-point Likert scale ranging from 1 – very unsatisfied, to 5 – very satisfied. Vertical bars indicate the % of replies and are positioned such that positive responses are shown above 0% and negative responses below 0% (the center of the ‘Neutral’ condition). Health care professionals: Cat 1 N = 34, Cat 2 N = 25, Cat 3 N = 34. Parents/caregivers: Cat 1 N = 16, Cat 2 N = 4, Cat 3 N = 14). Post-hoc comparison between pairs of categories (per group) were Bonferroni corrected for number of comparisons, N = 3. * p < 0. 05, *** p < 0.001.

Themes that emerged from the additional open questions on experienced communication barriers and current AAC extend these findings. From the answers to these questions, we identified three themes related to the origin of the communication barriers:1) the individual (child with CP), 2) the AAC device, and 3) the (social) environment. Recurrent factors brought up by both parents/caregivers and health care professionals related to the individual are that visual impairments make it harder to control AAC devices such as eye trackers and that children with CP are easily overstimulated (*‘All our children have a visual impairment, which seriously hinders the use of communication tools’; ‘My son’s head fills up quickly’*). Problems with AAC devices were mainly raised by parents/caregivers and concerned issues such as complexity of the device, duration/costs of maintenance and the size of the devices. Pertaining the (social) environment, problems were raised mainly by the health care professionals and almost all related to communication partners underestimating the cognitive and communicative abilities of children and adolescents with CP.

Of the communication aids used by the children and adolescents cared for by the respondents, Cat 1 was the overall most selected option by both groups (Cat 1 subcategories selected in total 166 times for health care professionals and 54 times for parents/caregivers; multiple selections possible per respondent), followed by Cat 3 (111 times for health care professionals and 20 times for parents/caregivers) and Cat 2 (30 times for health care professionals and 4 times for parents/caregivers) **(Figure 2C)**. In both groups, most respondents reported the production of sounds and words (‘Sound/word production’) as the most frequent form of communication. Most health care professionals also reported communication through movement of hand and fingers (‘Hand/fingers movement’) or ‘Head movement’ **(Figure 2C)**. For Cat 3, ‘Eye-trackers’ and ‘Touchscreens’ were most frequent reported by both groups. Both health care professionals and parents/caregivers reported significantly higher levels of satisfaction for more sophisticated communication strategies/aids **(Figure 2D)**. More specifically, satisfaction for Cat 3 was significantly higher than for Cat 1 in both groups, and significantly higher than Cat 2 for health care professionals (Kruskal-Wallis test across 3 categories per group, p < 0.05; Post-hoc comparison between pairs of categories per group, Bonferroni corrected for N=3, p<0.05; Mann-Whitney test between groups within category, non-significant).

The final open question of this part of the survey asked about general wishes and needs for children and adolescents with CP regarding communication. For health care professionals, themes that emerged were customizability, functionality, education, environment, personalization, and accessibility. Customizability referred to wishes and needs about having more degrees of freedom in the design of AAC devices, such as ‘*choosing your own vocabulary’*. Functionality was about adding more functions to existing AAC devices, such as *‘more integration of indirect communication like mailing, gaming, WhatsApp, social media*’; *‘longer battery life’* and *‘usability in more weather conditions’*. Wishes and needs labeled as education related to expanding knowledge about CP amongst schools, parents/caregivers, health care professionals and the public. Environment referred to considering the social and physical environment when thinking about better communication for children and adolescents with CP (*‘There is still an assumption that some children cannot learn to communicate better because their learning ability is too limited’*). Personalization referred to designing devices that fit the abilities of a specific child or adolescent with CP rather than a one-size-fits all solution. Finally, accessibility referred to wishes and needs about more social issues such as costs, insurances, and equal access to AAC devices. For parents/caregivers the emerging themes were autonomy, communication, functionality, and environment. Autonomy concerned the wish or need to communicate without assistance. Wishes and needs around communication were more and faster communication (*‘Being able to communicate faster!’)*. Functionality again concerned adding functions such as integrating a gaming application. Finally, wishes and needs concerning the social and physical environment were raised by the parents/caregivers, focusing on *‘More patience of communication partner*, and wishing for *‘schools with special education where assistive communication is fully embedded in the education’*.

### 3.3 Interest in BCIs

In the last section of the survey, we asked participants questions regarding BCIs and in particular cBCIs. Half of the health care professionals (18/36) and about half of the parents/caregivers (9/19) were naïve to the concept of BCIs, whereas others had read about BCIs in books, scientific articles or news articles (10 and 6, respectively), or had seen BCIs in action on TV or in person (8 and 4, respectively, of which one parent tried a BCI themselves) **(Figure 3A)**. Only one parent reported a BCI had been applied to their child. Three health care professionals applied BCI on a person. Before this study, impression about BCIs was generally positive **(Figure 3B).** There was no significant difference in the opinions of both groups (Mann-Whitney test, ns).

**Figure 3.**
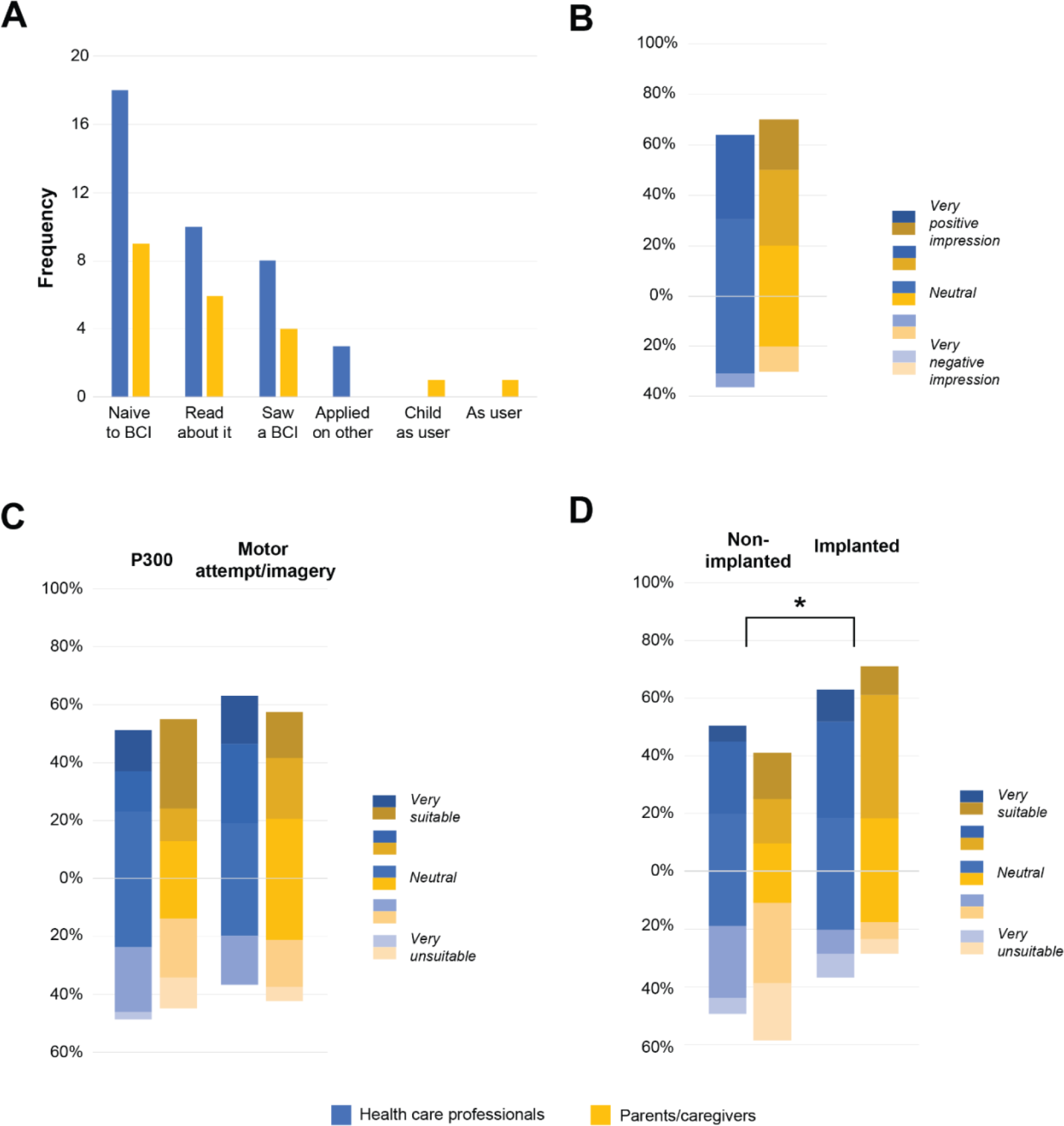
Interest in communication BCIs. (A-D) Health care professionals are indicated in blue and parents/caregivers in yellow. (A) Frequency of respondents with (no) prior knowledge or experience with BCIs before this survey. (B) Percentage of respondents who rated their impression of BCIs (if any previous experience) prior to this survey with a 5- point Likert scale (1 – very negative impression, to 5 – very positive impression; 18 health care professionals, 10 parents/caregivers). Mann-Whitney test between groups, non-significant. (C-D) Percentage of respondents who evaluated the suitability level for (C) each mental strategy (motor imagery/attempt or P300) for cBCI control and (D) different placements of electrodes (non-implanted or implanted), both using a 5-point Likert scale (1 – very unsuitable, to 5 – very suitable). Significant difference between non-implanted and implanted techniques when combining both groups (parents/caregivers and health care professionals) tested with Mann-Whitney test (*p < 0.05). (B-D) Vertical bars indicate % of replies and are positioned such that positive responses are shown above 0% and negative responses below 0% (the center of the ‘Neutral’ condition).

After introducing the concept of cBCIs, participants were asked about their opinion on the potential of cBCIs for the target population. Identified potential meanings and aims for both groups were autonomy, usability, communication, functionality, social, added value compared to other AAC devices, and self-expression. The health care professionals added cognition as an extra theme. Both groups indicated that cBCIs could make children and adolescents with CP more independent (autonomy), both in their ability to communicate and in having wider access to fulfilling their wishes and needs. As for the usability of cBCIs, doubts were raised for children and adolescents with CP and cognitive impairments, and, relatedly, excitement was raised for children and adolescents with CP with good cognition but severe motor impairments. One care professional expressed this sentiment when they said that *‘I think it could be a possibility for a select group of children. This will especially concern the group of severely affected children with dyskinetic CP. We also see many children with a GMFCS level V who do not have the cognitive ability to use BCI.’* Also other health care professionals expressed the worry that cBCI use requires good cognitive capacities, and may therefore not be suitable for children and adolescents with motor *and* cognitive impairment. For communication, the main meanings and aims for cBCIs were *‘easier and faster communication’*. Potential functions, besides communication, were entertainment and *‘doing school work’*. Social meanings and aims were about easier access to social interactions. Compared to other AAC devices, the main expressed hope was that it could be an improvement compared to eye trackers for children and adolescents who have a hard time controlling those (*‘Wow that would be great. Especially if eyes are not the right entrance’*). Finally, self-expression pertained to all the meanings and aims that have to do with children and adolescents with CP being able to better express their inner worlds, and, consequently, *‘be less underestimated’*.

Participants were subsequently asked to rate how suitable each presented mental strategy (visual stimuli or motor attempt) and recording method (non-implanted or implanted) would be for children and adolescents with quadCP (or their own child). Health care professionals and parents/caregivers had no significant preference for a mental strategy (Mann-Whitney test per group or with combined groups, ns), although health care professionals found motor imagery/attempt marginally more suitable than P300 **(Figure 3C)**. The main argument in favor of motor imagery/attempt rather than P300 was that P300 still requires visual attention, which might be difficult to achieve for children and adolescents with CP. The main argument in favor of P300 was that motor imagery/attempt requires users to imagine or attempt movements, which may be hard to accomplish for children and adolescents with CP who have no experience with generating movement. Notably, implanted electrodes were overall deemed more suitable than non-implanted ones **(Figure 3D)** (Mann-Whitney test between techniques with combined groups, p < 0.05; per group, ns). The main arguments for choosing implanted over non-implanted methods were 1) usability, 2) comfort, 3) independence, and 4) stigmatization. Both groups raised worries about the usability of non-implanted methods compared to implanted methods, as non-implanted requires the effort of putting on a cap with gel every time that someone uses the device. Moreover, the cap ‘*may have the disadvantage that they shift due to spastic movements or epilepsy*’, also making it less usable compared to implantable methods. Potential discomfort when wearing something on the head was also mentioned, for instance by a parent stating that their ‘*Son is very sensitive on his head*’, or a care professional that underscored this statement when they said that ‘*Some don’t tolerate anything on the head*’. Both groups stressed that non-implantable methods would make the child less independent about when they want to initiate communicating. A care professional noted that ‘*It requires the environment to apply the cap. That would mean the environment decides when the child may communicate’,* concluding that ‘*implanted electrodes seems much more suitable to me’*. Finally, both groups noted that non-implanted electrodes are visible to the outside world, which may lead to stigmatization for a group of individuals that is already stigmatized because of their disability. One care professional wrote that *‘Electrodes on the outside are vulnerable, difficult to apply to this target group and stigmatizing. An implant seems more suitable.’* And a parent wrote that *‘Appearance, how people look at him, is very important.’* The main argument against implanted methods were worries about the risks of surgery (*‘Messing with the brain sounds scary, including the risk!*’). Both groups also stressed that the right choice in this case may differ per individual.

## 4. Discussion

Knowledge about the needs and wishes regarding cBCIs for children and adolescents severely affected by CP is important for the development of cBCIs. The current study is the first to assess the perspectives on cBCIs of parents/caregivers and health care professionals of children and adolescents with quadCP. Parents/caregivers and health care professionals considered motor impairment the most frequent and impeding barrier to communication for children and adolescents with quadCP, were most satisfied with currently used high-tech compared to lower-tech AACs, and showed positive interest in the use of cBCIs, especially the implanted ones. They showed no clear preference for a mental cBCI control strategy, when choosing between motor imagery/attempt or visual stimuli such as P300.

### 4.1 Factors impairing communication

Parents/caregivers and health care professionals agreed on motor impairments being the most important cause of communication impairment in children and adolescents with quadCP. This is relevant for the consideration of cBCIs as a potential solution for this group, as these novel technologies aim to bypass lost motor function. In addition to motor function, health care professionals considered children’s cognition an important limiting factor for communication, whereas parents/caregivers found difficulties in communication with communication partners more important. These factors will be important to take into account in the development of cBCIs, and in the definition of the subgroups of children and adolescents with quadCP that may benefit the most.

### 4.2 Current AACs and wishes and needs regarding communication

Both parents/caregivers and health care professionals reported frequent use of no/low-tech and high-tech devices, but preferred currently-used high-tech AACs over lower-tech AACs. For AAC devices to be used in an effective way and to improve participation in everyday life of children and adolescents with quadCP it is important that the device meets the user’s needs **(Cockerill et al. 2014)**. It is therefore pivotal to consider the wishes and needs regarding communication of the user and their environment. In the current study, both parents/caregivers and health care professionals expressed their wish regarding communication to add more functions to existing AAC devices, for example the integration of a gaming application or indirect communication tools such as email, and to consider the social and physical environment. Furthermore, parents/caregivers expressed the need for faster communication than is possible with existing AAC devices plus the ability to self-initiate communication. Moreover, conventional AAC devices were identified with issues such as costs, complexity, and size of the device that had impact on the ease of communication. Some of these issues, such as costs, may not be solved by BCIs, and modifications of existing AAC devices may already serve individual’s needs. However, the reported factors will be essential to consider in the development of cBCI devices for those children and adolescents with quadCP who are unable to use conventional AAC and who may benefit from a cBCI.

### 4.3 Potential of BCIs for children and adolescents with quadCP

The concept of a cBCI appears to be of interest for parents/caregivers and health care professionals of children and adolescents with quadCP. Importantly, the use of cBCIs may be particularly valuable for a select group of children with CP who have age-appropriate cognitive and language comprehension abilities but severe motor impairments. Overall, both groups of respondents seemed somewhat more interested in implanted BCIs compared to non-implanted BCIs. This is in line with previous studies showing a positive attitude towards implanted BCIs of adults with ALS **(Blain-Moraes et al. 2012; Huggins, Wren, and Gruis 2011; Lahr et al. 2015; Liberati et al. 2015)** and spinal cord injury **(Huggins et al. 2015)**. The interest for implanted BCIs by individuals with motor disabilities appears to stem, among others, from the stigmatization associated with non-implanted BCIs, due to the visibility of an EEG cap. Another factor expressed by both ALS and spinal cord injury patients was the long setup time and associated suboptimal ease of use for non-implanted BCIs. This was also brought up by parents/caregivers and health care professionals in the current study. These arguments may be of extra importance to this young population that strives for autonomy, which is particularly complicated to accomplish by young people with severe CP **(Schmidt et al. 2020).**

Parents/caregivers and health care professionals did not show a strong preference for a mental strategy for BCI control (i.e., P300 or motor imagery/attempt). This result is not in line with earlier findings on caregivers of adults with locked-in syndrome (LIS), who seemed to prefer P300 over attempted or imagined or imagined hand movement (although adults with LIS themselves had the opposite opinion) **(Branco et al. 2021)**. Differences in the development of the motor impairment may explain this finding. Individuals with LIS and with CP differ in the sense that CP is a non-progressive neurodevelopmental disorder that manifests around birth or in the first year of life, whereas LIS often occurs later in life. As participants in the current study mentioned, and as previous BCI studies showed **(Daly et al. 2013; 2014)**, individuals with severe CP may have never performed a specific voluntary motor imagery/attempt, making BCI control that relies on that mental strategy difficult. Moreover, a different organization of the motor cortex after early brain injury may cause alterations in EEG activity, which can impact the effectivity of motor imagery/attempt to drive BCIs **(Daly et al. 2014; Jadavji et al. 2021)**. On the other hand, some individuals with CP may have difficulties to achieve visual attention, making the P300 a less suitable mental strategy for them. Thus, BCI control for children and adolescents with quadCP will be challenging and may need to be customized per individual.

Considering the high diversity within the group of children and adolescents with CP, in terms of severity and type of CP, due to variation in the nature and location of brain lesions **(Rosenbaum et al. 2007)**, it will be important to adapt the development of BCIs to individuals’ needs. The type of cBCI that suits best may not be the same across individuals, asking for a personalized approach. Such personalization may also be required at the level of the selection of BCI control features. A recent study by **Tou et al. (2023)** showed that individualized electrode selection in children with severe CP appears to improve the calibration accuracy of BCIs, while this was less the case for children with mild CP, and not beneficial for typically developing children.

### 4.4 Strengths and limitations

This is the first national study that included a large number of health care professionals working with children and adolescents with quadCP. They work with the target group in daily practice but can also approach the topic from a distance since they have no close personal relationship to them. This makes their opinion on cBCIs for children and adolescents with quadCP relevant for the development of cBCIs. Together with the responses from parents/caregivers, this study provides indications of the needs and wishes of this group. Whereas this study was designed and executed by a team that included CP patient representatives, it did not investigate the opinion on cBCIs of the children and adolescents with quadCP themselves. Their needs and wishes regarding cBCIs may differ from that of their parents/caregivers and health care professionals **(Blain-Moraes et al. 2012; Liberati et al. 2015; Branco et al. 2023)**. Moreover, their desire to use an assistive device such as a cBCI may depend on their perception of the advantages of cBCIs **(Hemmingsson, Lidström, and Nygård 2009).** We are planning a follow-up study on these questions with the young cBCI target population with CP. Yet, in this particular situation, it should be acknowledged that the opinion of health care professionals and parents/caregivers is relevant, since these groups typically have a decisive voice in the care for and support of children and adolescents with quadCP.

## 5. Conclusion

In this study, we investigated the needs and wishes regarding cBCIs as an AAC technology for children and adolescents with quadCP. We showed that parents/caregivers and health care professionals consider the use of cBCIs, especially implanted ones, as a promising AAC solution. Due to large variability within the group of children and adolescents with quadCP, a personalized approach for the development of cBCIs will be necessary. The current results are encouraging for cBCI developers, but assessing the opinion of children and adolescents with quadCP themselves is a crucial next step in the development of the most feasible cBCI system for use in everyday life.

### Data availability

Video animations (narrated in Dutch) used in this study are openly available in https://www.youtube.com/@umcuribs9505/videos.

### Ethical statement

The study was assessed by the medical research ethics committee of the Utrecht Medical University Center, who deemed this study exempt from the Dutch Medical Scientific Research Act (non-WMO).

## Acknowledgements

The authors thank all participants who took the time to participate in this research and Merel Horsmeier for designing and producing the animation videos. This research was supported by the Dutch Technology Foundation STW (PANDA project, grant 19072, MPB), the Kinderrevalidatiefonds Adriaanstichting/JFK Kinderfonds (OTTER project, grant 20210009, MJV), and the Gravitation program of the Dutch Ministry of Education, Culture, and Science and the Netherlands Organization for Scientific Research (ESDiT project, grant 024.004.031).

